# Impact of the Covid-19 pandemic on the mental health and wellbeing of adults with mental health conditions in the UK: A qualitative interview study

**DOI:** 10.1101/2020.12.01.20241067

**Authors:** Alexandra Burton, Alison McKinlay, Henry Aughterson, Daisy Fancourt

**Author notes:** Corresponding author: Dr Alexandra Burton, University College London, Department of Behavioural Science and Health, 1-19 Torrington Place, London, WC1E 7HB.

## Abstract

**Background:** People with mental health conditions have been identified as particularly vulnerable to poor mental health during the coronavirus disease 2019 (COVID-19) pandemic. However, why this population have faced these adverse effects, how they have experienced them and how they have coped remains under-explored.

**Aims:** To explore how the COVID-19 pandemic affected the mental health of people with existing mental health conditions, and to identify coping strategies for positive mental health.

**Methods:** Semi-structured qualitative interviews with 22 people with mental health conditions. Participants were purposively recruited via social media, study newsletters and third sector mental health organisations. Data were analysed using reflexive thematic analysis.

**Results:** Participants were aged 23-70 (mean age 43), predominantly female (59.1%) and of white ethnicity (68.2%). Fifty percent were unable to work due to illness and the most frequently reported mental health condition was depression. Five pandemic related factors contributed to deteriorating mental health: i) feeling safe but isolated at home ii) disruption to mental health services, iii) cancelled plans and changed routines iv) uncertainty and lack of control, v) rolling media coverage. Five coping strategies were identified for maintaining mental health: i) previous experience of adversity ii) social comparison and accountability iii) engaging in hobbies and activities, iv) staying connected with others, v) perceived social support.

**Conclusions:** Challenges were identified as a direct result of the pandemic and people with severe mental illnesses were particularly negatively affected. However, some found this period a time of respite, drew upon reserves of resilience and adapted their coping strategies to maintain positive wellbeing.

## Introduction

The coronavirus disease 2019 (COVID-19) pandemic caused widespread negative effects on mental health globally, with large-scale surveys from countries including the UK identifying increases in mental distress amongst general populations (Fancourt, Steptoe, & F., 2020; Niedzwiedz et al., 2020; Pierce et al., 2020). Having a pre-existing mental health condition has been identified as a risk factor for higher reported levels of depression and anxiety during this time (Iasevoli et al., 2020; Iob, Frank, Steptoe, & Fancourt, 2020; O’Connor et al., 2020) and concerns have been raised that people with existing mental health conditions may be at risk of longer-term effects such as relapse (Holmes et al., 2020).

In considering why this population has been adversely affected, a number of theories have been proposed including (i) higher concerns about the virus itself, such as COVID-19 related delusional beliefs and health anxieties (Johnson et al., 2020); (ii) a greater propensity to be affected by societal restrictions due to higher baseline risk of loneliness and social isolation (Holmes et al., 2020; Michalska da Rocha, Rhodes, Vasilopoulou, & Hutton, 2018; Wang et al., 2020; Yao, Chen, & Xu, 2020); and (iii) disruption in mental health service provision (Tromans et al., 2020) as well as reduced availability of informal support networks (Johnson et al., 2020). However, to date, research into triggers of poorer mental health during the pandemic amongst people with mental health conditions has been predominantly quantitative, focusing on pre-assumed hypotheses. Further, having an existing mental health condition may not always be a precursor to poorer experiences; feelings of resilience and greater acceptance by others have also been reported, alongside the use of coping strategies learnt from the prior self-management of mental health conditions (Sheridan Rains et al., 2020). These findings were however, predominantly taken from media reports and data obtained from organisational websites. There remains a clear research gap for understanding these broader mental health experiences and the management of these experiences during the pandemic from the perspectives of those with lived experience of mental health conditions.

The aim of this study was to i) identify the impact of self-isolation, social distancing measures and the pandemic overall on the mental health and wellbeing of people with mental health conditions, and ii) identify protective factors and coping strategies that were applied to support positive mental health.

## Materials and methods

This was a qualitative study, based on semi-structured interviews with people with mental health conditions in the UK. The study took a phenomenological approach, focusing on participants lived experiences of the pandemic and their subjective meanings of the changes brought about by the social distancing restrictions (Langdridge, 2008).

The study was approved by the University College London Research Ethics Committee, reference number 14895/005.

### Participant recruitment

Inclusion criteria were adults aged 18+ living in the UK with a self-reported diagnosis of one or more mental health conditions who were able to speak English for interview purposes. Recruitment occurred via social media, the COVID-19 Social Study website and newsletter, and a mail out from third sector organisations working with people with mental health conditions. Participants were purposively recruited to ensure diversity in gender, age, ethnicity, and mental health diagnosis. Potential participants were emailed a study information sheet and asked to contact the researcher if they had any questions about the study. Written informed consent was obtained from all participants prior to interviews being arranged. Participants also completed a demographics form (See Table 1).

**Table 1:**
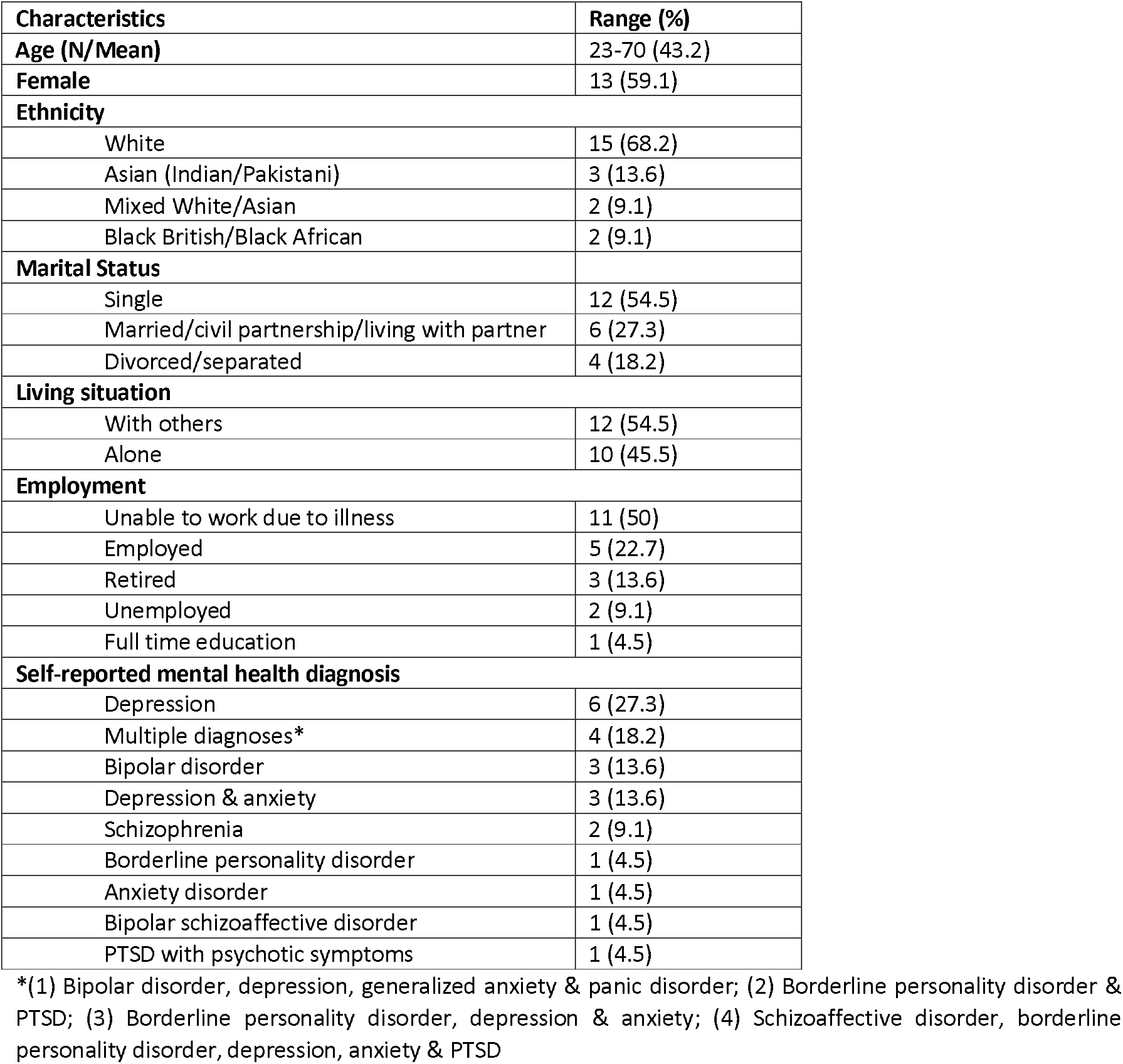
Participant characteristics.

### Data collection

Interviews followed a topic guide and were conducted by AB (mental health services researcher), AM (research psychologist), LB (sociologist) and HA (postgraduate research and medical student) remotely via video call, or telephone depending on participant access to technology and personal preference. Existing theories on behaviour change (Michie, van Stralen, & West, 2011), social integration and health (Berkman, Glass, Brissette, & Seeman, 2000), and health, stress and coping (Eriksson, 2017) guided topic guide development alongside emerging findings from COVID-19 Social Study survey participants and feedback from patients and the public via the MARCH Mental Health Research Network, both of which informed the inclusion of specific questions about mental health and protective activities. Questions explored i) participant adherence to social distancing guidelines, ii) changes to social lives, iii) impact of the pandemic on mental health, and iv) worries about the future. Example topic guide questions are listed in Figure 1. For the full topic guide please refer to Supplementary material File1_Topic_Guide.

**Figure 1.**
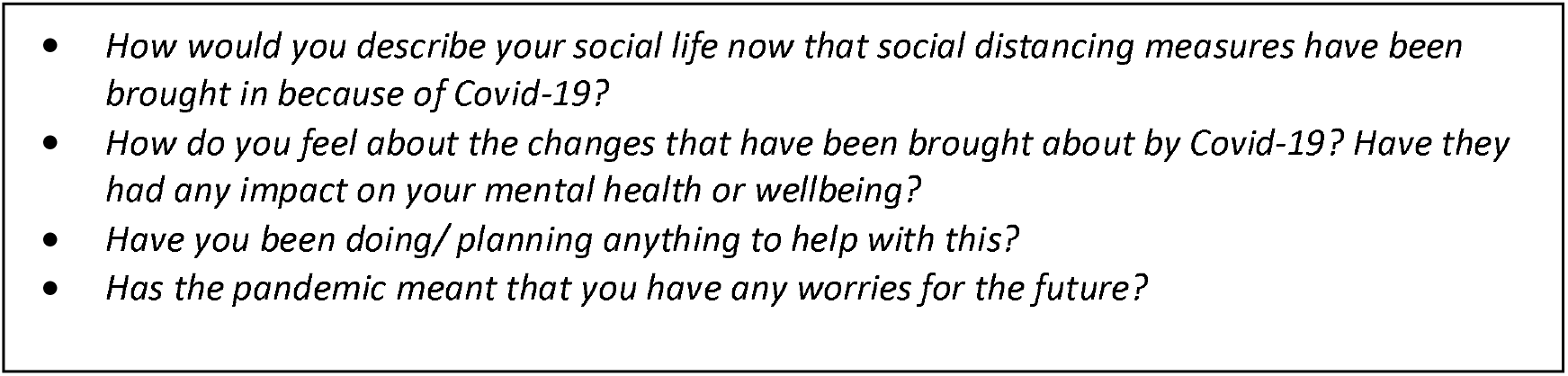
Topic guide question examples.

Interview length averaged 56 minutes (range = 25-81 minutes) and participants were offered a £10 voucher to thank them for their time. Interviews were audio recorded and transcribed verbatim by an external transcription company. Transcripts were checked for accuracy and anonymised by the lead researcher (AB) before analysis. Data collection continued until theoretical saturation, whereby no further concepts were discussed by interviewees (Braun & Clarke, 2006).

### Data analysis

Data were analysed using reflexive thematic analysis (Braun & Clarke, 2006; Braun, Clarke, & Hayfield, 2019). Transcripts were entered into Nvivo 12 software for analysis (QSR International Pty Ltd. NVivo, 2020) and familiarisation with data was achieved by reading through each transcript before coding commenced. A deductive analytical approach was initially taken with the development of a preliminary coding framework using concepts from the interview questions, including codes related to mental health, social isolation, coping strategies and barriers to engagement in protective activities. This was followed by inductive coding whereby new codes were generated from participant accounts and added to the coding framework. The coding framework was applied to the transcripts using line-by-line coding. Two researchers (AB, AM) independently and systematically coded three (14%) of the transcripts and discussed the generated codes and emerging themes. <Author1> continued coding the data using the revised framework, adding new codes to the framework as more transcripts became available and until no new codes were identified. Codes were then organised into themes corresponding to the research questions. The research team met weekly to review and discuss codes, themes and developing findings and to approve the final results and report.

## Results

22 participants were interviewed between May and August 2020. Participants were 23-70 years old (mean age 43) and were predominantly female (59.1%) and white (68.2%). Just over half were single and just under half lived alone. Fifty percent were unable to work due to illness and the most prevalent mental health condition was depression. Participant characteristics are presented in Table 1.

### Themes

Five pandemic-related factors contributed to a reported deterioration in mental health. Five coping strategies and protective factors were also identified (Figure 2). Themes, sub themes and corresponding participant quotes are presented below. For additional participant quotes illustrating each sub theme please refer to Supplementary table1_Additional quotes. Where appropriate we have included contradictory data alongside dominant accounts to capture subtle nuances in experiences.

**Figure 2.**
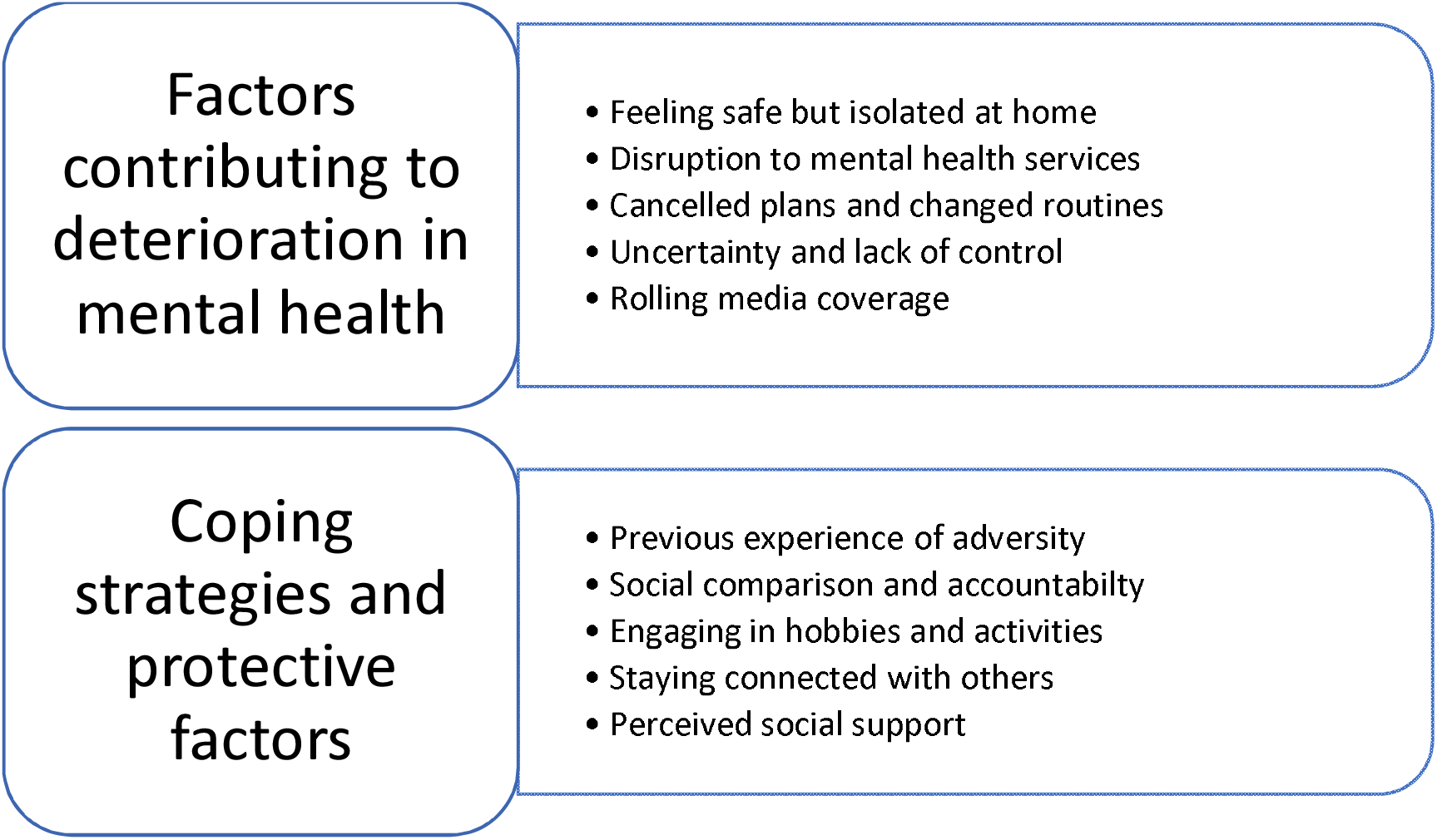
Key themes corresponding to the research questions.

### Factors contributing to deterioration in mental health

#### Feeling safe but isolated at home

Leaving the house was a cause of anxiety for many participants who were worried about contaminating their homes, about other people getting too close to them, and ultimately catching the virus.

> *I have to leave the flat to get my post or to take rubbish out. I’ve noticed an increase in panic attack lead up symptoms if that makes sense*… *happening a lot more than it has done for a very long time. (ID09)*.

Some participants reported feeling happy at home and limiting contact with the outside world, however they acknowledged that while this felt safe, being isolated for prolonged periods was detrimental to their mental health. For some, this felt like a step backwards in their recovery.

> *It was nice not to have to go out, it was like I had an excuse*…*Lockdown is a good thing for me in the way that I could isolate*…*but on the other hand, isolating like that is bad for me because that’s how I get ill (ID25)*.

Others described prolonged periods spent indoors as challenging because of the lack of physical contact with others. Those who lived alone or who relied on connecting with friends and family as a coping strategy to protect their mental health were particularly affected.

> *I’m a very social person and probably seeing people is the thing that makes me less anxious. Those are the things that make me feel better. So obviously I couldn’t do any of them, and I think the more time I spend on my own, the more anxious I am and the more my mental health suffers (ID06)*.

#### Disruption to mental health services

Some participants described difficulties accessing mental health services and experienced a perceived or actual lack of service provision during this time.

> *In crisis, it’s the local A&E for me*…*and obviously that’s not on at the moment either, because my local hospital is the local infectious diseases one and they’ve shut down completely, apart from COVID (ID09)*.

Those with severe mental health conditions experienced a withdrawal of community services at the start of the lockdown, which led to reduced practical and emotional support, greater feelings of isolation and a decline in mental health.

> *During lockdown (support worker) stopped coming*… *It wasn’t explained to me, I assumed that during lockdown I would receive the same kind of support and it wouldn’t be affected, but it was unexpected, and it put a lot of pressure, a lot of turmoil, on top of me, living with schizophrenia*… *you’re not managing (ID19)*.

For those who were able to book virtual consultations with health professionals, these were often experienced as inferior to face-to-face appointments.

> *It’s not the same because you can’t get across your emotions and what’s going on in my head, very well. Not that I sit there and cry or anything but it’s just not the same (ID07)*.

A small number of participants described the importance of third sector mental health services. However, for some, services had closed or moved online and were no longer perceived as helpful.

> *I used to use a mental health service called <anonymised>. And that’s probably the thing that hit me the hardest was not being able to go there*…*I’m really hoping that it doesn’t disappear as a result of this (ID15)*.

A minority of participants expressed concerns about the future provision of mental health services and the impact the pandemic might have on their ability to access support longer term.

> *And, I’m concerned, it is so stretched now getting any support from mental health services, and I’m frightened it’s going to get even worse now with the increased number of people who will now be suffering from mental health problems; that I’m not going to get any support at all (ID12)*.

#### Cancelled plans and changed routines

Disruption to holidays and cancelled celebrations and events led to frustration and disappointment among many participants, with some describing feeling in limbo or trapped.

> *I think this feeling of entrapment is, just the biggest fear for me not being able to run away from things*… *not being able to get away from something like [City] can be really claustrophobic (ID11)*.

Uncertainty around when social activities could resume safely was a cause of concern, with some participants unsure as to how they could feel comfortable in close proximity to others within larger social spaces. There was a sense among participants that social interactions would need to be conducted differently in the future.

> *I’ve got a thing on my phone what I was doing this time last year, and I was at a folk festival in the park*…*and there were pictures of people all very close together*…*and that just seems strange now*…*like there’s something wrong with it (ID18)*.

Other participants, however, described the importance of making plans even amidst the uncertainty, to have something to look forward to that resembled normality.

> *They’ve cancelled <community festival> this year now. So, I might be looking at next year now. That’s one thing I’ve got for the future (ID20)*.

As well as cancelled plans, participants described disruption to daily routines as a trigger for low mood and anxiety.

> *It’s hard to stick to a routine, so just keep occupied, try and keep engage in the activity. But I do feel disengage, little interest. So, I might feel forget about today, and work towards another day, towards tomorrow (ID19)*.

Some participants found that the lack of activities led them to spend more time ruminating over negative thoughts. Without the distraction of leaving the house to break up the days, these thoughts could become overwhelming.

> *The problems that I would feel have been more brought about by the time to think, which is not always productive*…*because we’ve not been going out and not diverting thoughts away from those sort of situations (ID13)*.
>
> *I’ve always been a self-harmer, I haven’t done it since last year sometime when I was in hospital. And one thing I have noticed about symptoms that have happened during lockdown, I didn’t have any for a really long time before lockdown and I feel like lockdown made me want to do that. Not like there’s some reason but it’s given me the time or the isolation to dwell on things and be in a different headspace (ID25)*.

Others highlighted the importance of maintaining or developing new routines, either by adapting ways of working and communicating, or with support from others.

> *I’ve spoken to my psychologist from DBT days and she and I have worked out a plan to go forward, for the immediate future. That involves day-to-day routine*…*just trying to get a routine going again (ID04)*.

#### Uncertainty and lack of control

Many participants described the ongoing uncertainty of the pandemic and changing restrictions as negatively affecting their mental health. In the lead up to lockdown, the speed in which changes were imposed were experienced as deeply unsettling, with little time to say goodbye to friends or adapt to new ways of living and working.

> *There was such a panic, for me, because everybody was going back home, and didn’t let me know, like I didn’t have much contact with them in the pandemic, when we were first told*.*it was an evolving situation. It was quite concerning for me*…*I wasn’t even able to say goodbye (ID17)*.

Participants described feeling a lack of control over the changes that were being made to their daily lives. At times, this felt *“overwhelming”* due to *“not knowing”* how long restrictions would be in place with a feeling that *“nobody really has the answers”*. For some, however, the feelings of uncertainty transformed into a renewed acceptance and a welcome relinquishing of control.

> *I think it’s the fear of the unknown, so initially, the whole boat is put off course, and you can’t see where you’re going and what’s going to happen*…*I certainly don’t react well to change. Once that becomes the accepted norm for yourself*…*you can relax into it. It’s really surprised me (ID02)*.

Others feared the end of restrictions because they had become accustomed to a *“new normal”*, one that they had learned to adapt to and feel in control of.

> *Strangely, there’s part of me that doesn’t want it to end*…*It’s been a really stressful, horrible time but there’s also a part of me that when I hear about the potential of it ending, I feel nervous, scared (ID10)*.

#### Rolling media coverage

A number of participants described the rolling coverage of the pandemic as overwhelming. Some described difficulties avoiding the news, even though this was detrimental to their mental health, while others actively limited engagement with media to protect their mental health.

> *I try not to spend much time on the news at times, because it can be really, really upsetting (ID14)*.

For some participants with severe mental health problems, the coverage of the pandemic manifested itself within their symptoms.

> *I did get a hallucination of COVID infested bats flying through my window in a giant cloak*…*It always seems to be when we’ve had one of the big talks on the daily briefings on the TV. When something major’s happened with the lockdown that seems to trigger my bad hallucinations (ID12)*.

## COPING STRATEGIES AND PROTECTIVE FACTORS

### Previous experience of adversity

Participants described drawing on experiences of trauma and turmoil in their lives to make sense of the pandemic and revisited previously effective coping strategies.

> *Having a very turbulent childhood has made me very good at dealing with turbulence. So*…*stuff like the pandemic, with everything being really strange and different actually is a good thing for me because it makes me flip my head and go, okay, what can I do to make this better for me? (ID11)*.

Some reflected on their previous experiences of social isolation to help cope with the restrictions.

> *I thought oh God, can I cope with this for a long time? Then I remembered that I’d spent nine years when I didn’t go out much, and I thought yes I can manage it. I drew on that to think the things that I shouldn’t do and the things that I should do. That experience helped me (ID18)*.

### Social comparison and accountability

Some participants felt liberated by the pandemic restrictions and experienced a reduced sense of accountability to and judgement from others and less pressure to conform to social norms.

> *I have this time issue, like a race against everybody else, so I think lockdown gave me that feeling of, we’re all on the same platform right now; nobody’s doing anything*…*I know I’m not missing out on anything, but I’ve felt like I’m on an even platform with all my peers again (ID08)*.

A minority of participants however experienced the lockdown as a time for reflection on missed opportunities, comparing their lives against the achievements of their peers.

> *Lockdown has suddenly made it occur to me that I should have set milestones by now. Because I spent so long in hospital during my life, I haven’t hit the same milestones that my friends or my sister’s reached. The lockdown’s got me worried about my future plans*…*and uncertainty in what they are (ID25)*.

A number of participants described a renewed sense of understanding from friends and family who now empathised with the impact that having a mental health problem had on participants’ daily lives. This stemmed primarily from conversations about health anxieties and social isolation.

> *Quite a few of my friends didn’t really have that problem in the first place, and now they get it, feeling anxious about meeting people, feeling like you’re missing out, in a way. I feel like there’s been a lot more understanding, and that’s been a positive thing (ID16)*.

### Engagement in hobbies and activities

Most participants described engaging in social activities at home. For some, this was a chance to increase time spent on existing hobbies, while others discovered new interests.

> *We’ve been playing a lot of box games and I do a lot of cross-stitching, which is very mindful. I’m trying to keep my mind occupied so it doesn’t drift down into the abyss, as it were (ID07)*.

Not all participants were able to engage in activities that usually protected their mental health, for example, accessing community facilities such as museums, theatres or libraries.

> *A friend was saying to me on the phone, you don’t miss it that much because you weren’t doing much anyway*…*well yes, but me getting out once when I did*…*Getting to a museum, getting to a gallery, getting to a library was really important (ID09)*.

### Connecting with others

Spending quality time with family and connecting with friends and social groups online were important protective activities. Some participants experienced a renewed appreciation of friendships, particularly as online socialising required extra effort or patience.

> *That’s the one thing that we can’t do without. It’s the one thing that has been critical to the way that I’ve coped and just reinforcing because we’ve had to do things in a different way, I guess it’s made me much more grateful for those relationships because it’s had to be done differently (ID23)*.

For others however, social networks reliant on face-to-face contact had collapsed, or virtual ways of communicating were a poor substitute for meeting in person and contributed to a worsening of mental health.

> *I don’t have any sense of identity at the moment. I don’t really know who I am. Because I haven’t had much face-to-face contact with people, I don’t really get a sense of being real. Speaking through Zoom and WhatsApp and so on is a bit robotic (ID04)*.

### Perceived social support

Participants described the importance of knowing there was someone they could turn to if they needed emotional support. For some, support from family and friends was important for preventing relapse and identified as a necessary substitute in the absence of formal services.

> *The nurse, even after I mentioned I was struggling at the beginning of lockdown, never asked me again if I was okay or what support I’d got. It was just fortunate that I happened to have the support outside of the health service (ID23)*.

Participants had mixed perceptions about the practical support available to them. For some, family and neighbours were available to help with shopping for essentials, however others did not feel they needed support or felt too guilty to ask for help.

> *I made the decision that with not having a close family member there was no way I could ask a friend to do it, because I knew that if she contracted the illness I would feel guilty, I wouldn’t know if she’d got it through doing my shopping (ID22)*.

For some, a breakdown in support networks during the pandemic contributed to a deterioration in mental health.

> *I did all the shopping and everything myself. The eldest daughter used to help me to take medication but then we fell out and that all went out the window and I ended up taking that overdose because no one’s checking on me (ID12)*.

Being able to provide support to people in the community was an important protective factor for participants’ own mental health. Participants reported a need to feel useful during a time when social interactions were limited and others were in need.

> *A lady who lives at the end of the road did need (medication) picking up*…. *I found out she was self-isolating, but not considered high risk by the government and she was having trouble getting food*…*For two weeks (the government) sent (food packages) automatically so I took the boxes down to her. I felt really nice about being able to help her (ID16)*.

But, when participants were unable to fulfil these support roles due to the social distancing restrictions or because they were scared of infection, this resulted in guilt and frustration.

> *I’ve tried to do what I can to help other people while staying in, which has been hugely challenging, and quite stressful. Because you can’t just rush down to your friend when they feel ill, and that’s been really hard, to know that your friends are not well, and not just to be able to go and knock on their door, give them a card, give them a hug, or do anything at all (ID05)*.

## Discussion

Our study found that people with mental health conditions were experiencing particular challenges during the COVID-19 pandemic. Feeling isolated at home, reduced access to mental health services and the ongoing uncertainty surrounding imposed restrictions all contributed to a deterioration in mental health. For those with severe mental illnesses this was a particularly difficult time, presenting unique triggers for self-harm and COVID-19 related hallucinations. However, negative psychological experiences were not always sustained beyond the initial lockdown period or indeed uniform across participant accounts; some found this period a time of respite or were able to use previous experiences of adversity to draw upon successful coping strategies. Others experienced less social pressure and felt less socially excluded due to empathy from others. Engagement in hobbies, connecting with loved ones and perceived availability of social support were identified as important protective factors.

### Factors contributing to deterioration in mental health

Triggers of poorer mental health identified in our study echo findings from previous research on risk factors for mental illness such as social isolation (Michalska da Rocha et al., 2018; Wang et al., 2020), and findings that people with mental health conditions had a five times higher risk of severe loneliness during the pandemic (Bu, Steptoe, & Fancourt, 2020). Our findings highlight how the pandemic exacerbated these triggers and led to increased uncertainty and not feeling in control. Participants acknowledged a conflict between the importance of maintaining social distancing and isolating at home to feel safe, and the adverse consequences of social isolation on their mental health. Detrimental effects on mental health linked to social distancing restrictions could be long lasting and have been observed in previous pandemics up to six-months after the end of quarantine in people with mental health conditions (Jeong et al., 2016). Other triggers of poorer mental health included disrupted access to mental health services, also identified in a study by Gillard et al, 2021. Indeed, a reduction in referrals to mental health services and an increased use of emergency psychiatric services were reported during the pandemic (Mustafa, 2020). Further, whilst prior to the pandemic, the provision of mental health telecare was broadly supported by health professionals as an adjunct to face-to-face care (Langarizadeh et al., 2017), participants in our study identified the lack of physical presence of the clinician as problematic.

### Coping strategies and protective factors

In corroboration with previous research (Gillard et al., 2021), we found some reports of positive mental health outcomes from the pandemic. Shared experiences of social isolation and health-related anxieties led to a “normalisation” of emotional responses that may usually be discouraged or labelled as symptoms for some participants (Watson, 2019). Further, some participants found they were able to adapt and form new routines, engage in meaningful activities and draw upon previous adversity and isolation to actively cope. Our study also found that perceived social support and connecting with family and friends were important buffers against relapse in line with previous research (Wang et al., 2018). But these benefits required participants to have access to such support and where it was lacking these psychological benefits were not felt.

### Limitations

We relied on people self-identifying for the study, which may have resulted in people taking part who were particularly motivated or experiencing less severe mental health problems. Half the sample were, however, unable to work due to illness and many reported being in contact with mental health services. People were interviewed while they were experiencing initial lockdown restrictions and the earlier stages of the pandemic, rather than retrospectively, and the ongoing changes and prolonged uncertainty may have an impact on mental health and coping strategies in ways that have not been captured here.

### Implications

This research highlights that many individuals with existing mental health conditions experienced poor mental health during the COVID-19 pandemic. However, these experiences were nuanced: certain factors appeared to confer psychological support and mitigate worse experiences. Future work should explore the changing needs of people’s experiences as the pandemic continues, the longer-term support that people with mental health conditions may need and how best to provide support. Such work may require adaptation and tailoring of existing services and interventions to comply with social distancing restrictions, while remaining acceptable and accessible to those who have struggled with virtual services and support.

## Data Availability

The data are not publicly available due to their containing information that could compromise the privacy of research participants.

## Acknowledgements

We would like to thank Dr Louise Baxter for support with recruitment and interviews and Dr Anna Roberts, Dr Tom May, Joanna Dawes and Katey Warran who provided weekly feedback on emerging findings. The research team are also grateful for the support of a number of organisations with their recruitment efforts including the McPin Foundation, Alzheimer’s Society, Arts Beyond Belief, Taraki and MQ, and to all of the participants who took part in the study.

## Author Contribution

DF conceived the study and DF & AB designed the study. AB, AM & HA collected data for the study, analysed and interpreted the data. AB wrote the first draft. All authors provided critical revisions, read and approved the submitted manuscript. All authors had full access to the data in the study and can take responsibility for the integrity and accuracy of the data.

